# Structural and Functional Characterization of the Aorta in Hypertrophic Obstructive Cardiomyopathy

**DOI:** 10.1101/2023.05.23.23290086

**Authors:** AM. Ibrahim, N. Latif, M. Roshdy, P. Sarathchandra, M. Hosny, A. Elsawy, S. Hekal, A. Attia, W. Elmozy, A. Elaithy, A. Elguindy, A. Afifi, Y. Aguib, M. Yacoub

## Abstract

**Background and aims:** Changes in the phenotype and genotype in Hypertrophic Obstructive Cardiomyopathy (HOCM) are thought to involve the myocardium as well as extracardiac tissues. The extent and significance of extra-myocardial changes has not been adequately studied. We here describe the structural and functional changes in the ascending aorta of HOCM patients.

**Methods:** Changes in the aortic wall were studied in a cohort of 102 consecutive HOCM patients undergoing myectomy, and 10 normal controls. Biopsies were examined histologically, immunohistochemically and by electron microscopy. Changes in protein expression were quantified using morphometry and western blotting. Pulse wave velocity (PWV) was measured using Cardiac Magnetic Resonance (CMR), in 86 HCM patients compared to 166 age-matched normal controls.

**Results:** In HCM, the number of medial lamellar units (MLU) was significantly decreased, associated with an increase in the interlamellar distance and a preserved thickness of the aortic wall, as compared to controls. Electron microscopy showed an altered lamellar structure with disorientation of elastin fibers from the circumferential direction. There was an altered composition and orientation of smooth muscle cells. In addition, there was a significant decrease in alpha-smooth muscle actin, smooth muscle myosin, smooth muscle 22 and integrin beta1, and a significant increase in calponin and caspase3. Fibulins 1, 2 and 5, had a reduced expression in HOCM aortic biopsies. Functionally, PWV was significantly higher in HOCM patients compared to healthy controls.

**Conclusion:** In HOCM patients, specific molecular and structural changes in the composition and organisation of the arterial wall have been identified. This was associated with increased stiffness of the arterial wall.

**Translational Perspective:** This study sheds light for the first time on the altered lamellar organization in the aorta of Hypertrophic Obstructive Cardiomyopathy (HOCM), in addition to the Smooth muscle cells and Extracellular Matrix abnormalities, to explain the increased wall stiffness associated with patients clinical phenotype. The data provide insights on extra-myocardial targets that can have potential value for risk stratification and personalized therapeutics for HOCM patients.

## Introduction

Hypertrophic cardiomyopathy (HCM) is an inherited cardiac disorder affecting more than 1:500 in the general population ^1–3^. HCM has always been defined with various myocardial changes and concentrating on disease-causing mutations related to sarcomeric genes ^4–6^. However, recent evidence has shown that HCM is heterogenous both at genotype and phenotype levels and involves many pathways and extra-myocardial tissues, such as the aorta ^7,8^.

Changes in the aortic stiffness have been reported in HCM ^9,10^ and correlated to exercise capacity ^11^. To date, the structural changes responsible for abnormalities in the aortic wall have not been studied. The wall of the aorta is highly elastic and dynamic which facilitates its function and response to the hemodynamic environment ^12–14^. One of the major players that determine the arterial wall stiffness, and the physical properties of the aorta, is the medial elastic lamellar number and composition, which was first described by Glagov and colleagues ^15–17^. This observation has since been the subject of many studies focusing on the medial lamellar unit (MLU) organization in physiological and pathological conditions ^18,19^, which sometimes yielded contradictory results due to the lack of adequate imaging techniques that can determine the 3D structure of the MLU. A recent 3D modeling and imaging of MLU clarified their exact composition and the functional interaction between the constituting elements. ^19^. The interlamellar space contains smooth muscle cells (SMCs) and different extracellular matrix (ECM) components, which play a critical role in determining the physical properties of the aortic wall, including stiffness ^20–23^. Elastin and collagen are the major components of the aortic wall ECM, however, other matrix proteins such as fibrillin, fibulins (FBLNs), and matrix metalloproteinases also play an important function in the aortic wall structure ^21^.

We here study in detail the number of elastic lamellae, and the width and composition of the interlamellar space, particularly in relation to the changes in ECM components as well as SMCs.

## Methods

### Patients samples

In all, 268 subjects were clinically examined (102 HCM and 166 controls [from the ECCO-GEN study^24^](**Table 1** and **Supplementary data**). Written informed consent was obtained from HCM patients and healthy subjects prior to their inclusion in the study [(20130405MYFAHC_CMR_20130330) and (20151125MYFAHC_Hvol_20161027), respectively]. Control aortic biopsies were acquired from the Magdi Yacoub Institute UK, upon a material transfer agreement [Royal Brompton hospital ethics review board / Brompton and Harefield trust ethics committee (REC approval 10/H0724/18)]. The study is abiding by all the standards of the Declaration of Helsinki.

**Table 1:** Clinical characteristics of Hypertrophic Cardiomyopathy (HCM) and Healthy volunteers [HVol] cohorts.

| Clinical Parameters | HCM (n = 102) | Hvol (n=166) |
| --- | --- | --- |
| Gender, Male n (%) | 74 (72) | 93 (56) |
| Age, years | 39.4 ± 14.3 | 32.8 (25–36) |
| Body surface area BSA. m<sup>2</sup> | 1.9 (1.71–2.1) | 1.8 (1.7–2.0) |
| Body mass index | 28.7 ± 6.2 | 26.4 (22.8–29.2) |
| Dyspnea n (%)<br>New York Heart Association (NYHA) class | 95 (93)<br>Class I (n = 4)<br>Class II (n = 31)<br>Class III (n = 51)<br>Class IV (n = 9) | 0(0) |
| Angina n (%) | 51 (0.5) | 0(0) |
| Syncope | 24(23) | 0(0) |
| Max Septal wall thickness (cm) | 2.4 (2– 2.8) * | NA |
| LVOTO resting gradient (mmHg) | 103 (80–128) | NA |
| Left ventricular mass by CMR (gm) | 204.5 (163.8–298.5) * | 85.19 (62–106) |
| Left ventricular end systolic volume by CMR (ml) | 19 (14–27) <sup>α</sup> | 51.9 (39–61) |
| Left ventricular end diastolic volume by CMR (ml) | 78 (67–93) <sup>α</sup> | 134.58 (113–154) |
| Ejection Fraction% by CMR | 74.9 ± 8.21<sup>α</sup> | 63.4 (59–67) |
| Diastolic dysfunction n(%)<br>Diastolic dysfunction Grade | 102(100)<br>Class 1 (n = 93)<br>Class 2 (n = 7)<br>Class 3 (n = 2) | 0(0) |
| Cardiac Troponin I (ng/mL) | 0.014 (0.009–0.03) <sup>¥</sup> | 0.003 (0.0001–0.002) <sup>∞</sup> |

\*The data are available for 91 patients only, <sup>α</sup> the data are available for 92 patients only, <sup>¥</sup> the data are available for 75 patients only, <sup>∞</sup> the data are available for 158 patients only, <sup>Σ</sup> the data are available for 86 patients only. Data are presented as mean ± SD for normally distributed continuous variables, as median and interquartile range for non-normally distributed continuous variables, and as percentages (%) for categorical variables.

### Tissue processing and histological staining

Thoracic aortic biopsies were collected, processed and prepared for staining as described before ^25^. Briefly, for fibrous collagen staining, slides were incubated in picrosirius red stain (Abcam) for 1 hr and washed in 0.5% acetic acid for differentiation. Sections were dehydrated with increasing concentrations of ethanol, cleared with two changes of xylene (Sigma) and finally mounted with DPX (Sigma). Slides were scanned and analyzed with a slide scanner (Zeiss-Axioscan).

### Immunohistochemistry and immunofluorescence

Section preparation was performed as described before ^25,26^. All antibodies were diluted using blocking solution; α-SMA 1:300 (Thermo Scientific™, # MS-113-P, SMM 1:200 (Abcam, ab 133567), SM22 1:200 (abcam, ab14106) calponin 1:200 (Dako, M3556), ITGβ1 1:150 (Abcam, #ab52971), Caspase3 1:50 (Abcam, #ab13847) FBLN2 1:200 (Thermo Scientific™, #PA5-21640), FBLN1 1:200 (Thermo Scientific™, #MA5-24598) and FBLN5 1:200 (Thermo Scientific™, #MA5-15395). Sections were incubated for 1 hr with HRP-goat secondary antibody (Dako) and washed thrice prior to staining with DAB plus Chromogen (Dako) for 1 min. For fluorescence reactions, washed sections were incubated with secondary antibodies conjugated with fluorophores, (Goat Anti-Mouse IgG H&L Alexa Fluor® 594) (abcam, ab150116) and (Goat Anti-Rabbit IgG H&L Alexa Fluor® 488) (Abcam, #ab150077) at a 1:500 dilution, incubated in the dark for 1 hr at RT, and then mounted with DAPI-containing mounting media (Invitrogen™, Carlsbad, CA, USA, #P36962). The number of lamellae and interlamellar distance were measured using Glagov’s methodology ^16,27^.

### Protein isolation and immunoblotting

Tissue biopsies were lysed and prepared for separation as described before ^25,28^. Briefly, 5 μg of protein per sample was separated and then transferred from the SDS polyacrylamide gel onto Whatman® Protran® Nitrocellulose Transfer Membrane (0.2 μm) (Sigma) using a Biorad transfer module, protein transfer buffer (1X NuPage transfer buffer, 10% methanol in dH2O). The membrane was incubated for 30 min at RT in 5% Marvel Original dried skimmed milk blocking solution (in 1X PBS Tween-20 wash buffer). The membrane was then incubated for 2 hrs at RT with primary antibody (GAPDH (Cell Signaling, 2118S) at 1:500, FBLN2 at 1:1000, FBLN5 at 1:100 and FBLN1 at 1:1000), washed thrice for 15 min, incubated for 1 hr at RT with horseradish peroxidase (HRP)-labelled secondary antibody (Life Technologies) and washed thrice for 15 min. Reaction with HRP was carried out using ECL Western blotting detection reagents (Life Technologies) and the signal was detected using an Amersham Imager 600.

### Electron microscopy

2 mm^2^ samples were processed as described before ^29^. Specimens were embedded in rubber molds, polymerised in the oven at 60°C. Ultra-thin sections (100nm) were collected on coated grids, post-stained with 2% aqueous Uranyl acetate and Reynolds lead citrate and viewed using JEOL 1200 transmission electron microscope. Digital micrographs were taken using a Gatan digital camera.

### Pulse wave velocity

Cardiac magnetic resonance (CMR) was performed with a 1.5 T MRI machine (Magnetom Aera - Siemens Healthineers, Erlanger, Germany). Pulse wave velocity (PWV) was calculated using Segment software (Medviso AB, Lund, Sweden), as per the formula; **PWV =Δ D / ΔT (m/s)**, where ΔD was the aortic path length between the mid-aortic valve and mid-descending aorta, and ΔT was the time delay between the arrival of the foot of the pulse wave at these levels. (**Supplementary methods and Supplementary Figure 1 and 2**).

## Results

### 1. The number and organization of MLU in HOCM aortic walls

The interlamellar distance was significantly increased in HCM aortic walls when compared to controls, which was measured between two adjacent elastin fibres or two adjacent SMCs (α-SMA+/ITGβ1+) in the circumferential direction of the fibres. Additionally, the number of lamellae was significantly reduced in HCM-aortic walls. However, the overall thickness of the aortic wall remained unchanged in HOCM (**Figure 1B and 1C**).

**Figure 1:**
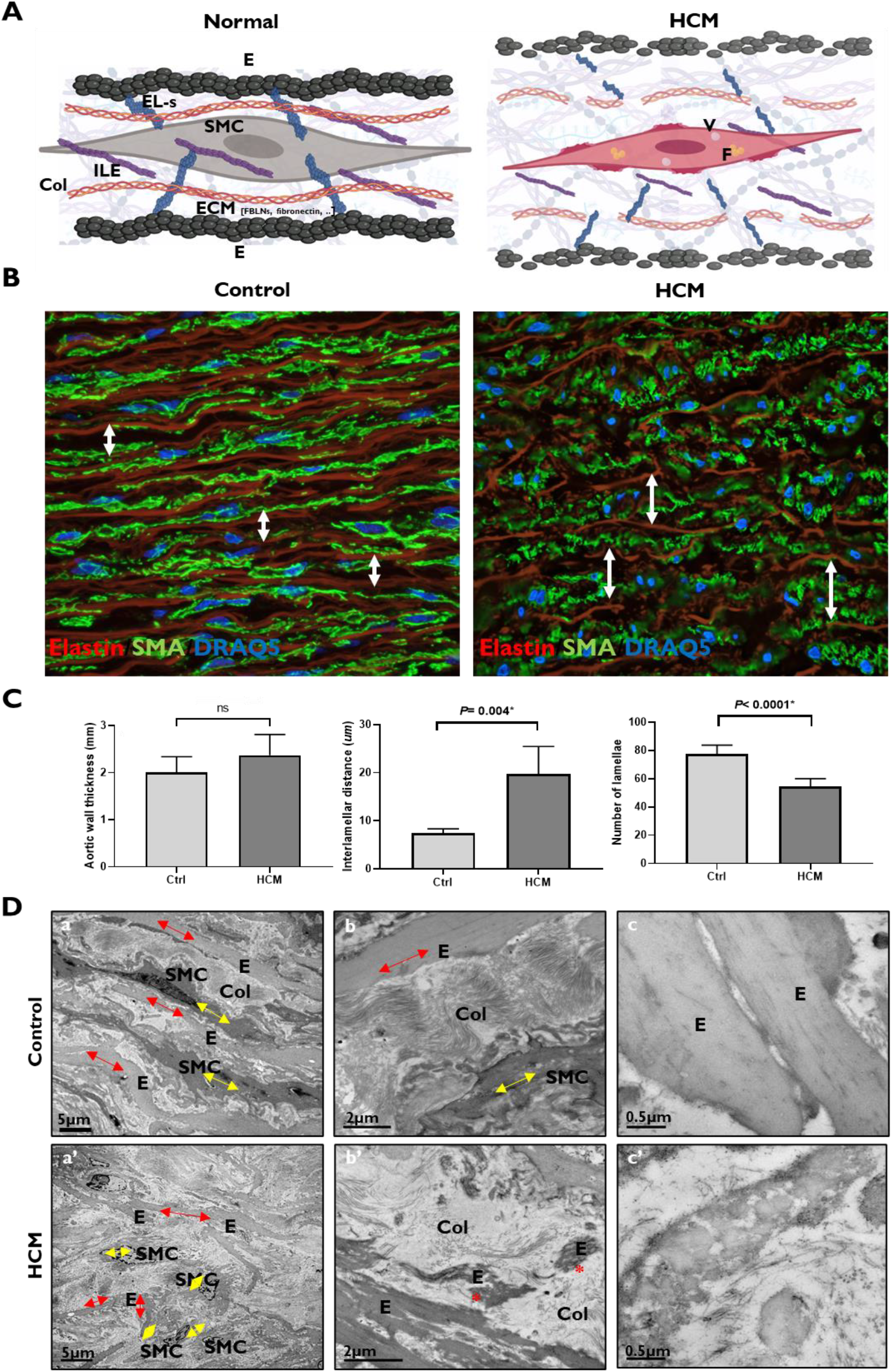
Lamellar Organization in HCM aortic wall. **A:** Schematic diagram shows the medial elastic lamella organization in the aortic wall [adapted from El-*Hamamsy and Yacoub, 2009, and O’Connell et al*., *2008*]. **EL:** elastin, **ILE:** interlamellar elastin, **EL-s:** elastin spurts, **COL:** collagen, **SMC:** smooth muscle cells, **ECM:** Extracellular Matrix, **V:** Vacuole and **F:** Fat globules. **B**: Confocal Microscopy shows the altered lamellar organization in HCM aortic walls, exhibited in the expression of SMA (green) in contrast to elastin fibres (maron). White arrows point to the interlamellar distance. **C**: Quantification of wall thickness, number of lamellae and the interlamellar distance in HCM (n=12) vs controls (n=3). **D**: Electron micrographs of control (a, b and c) (n=3) and HCM (a’, b’ and c’) (n=14) aortic walls. Asterix refers to radial elastin fragmentation a and a’ (magnification 5K, bar=5 μm), b and b’ (magnification 12K, bar=2μm), c and c’ (magnification 40K bar= 0.5μm).

Electron microscopy analysis of control aortic walls showed parallel circumferential organisation of lamellae and SMCs, with tightly packed interlamellar collagen in wave like formation (**Figure 1D-a, b and c**). Elastin fibers were thick, continuous lamellar sheets, with interlamellar elastin fibers and radial rods ^19^. At lower magnification, the HCM walls showed disorganisation of the lamellar unit associated with disorientation and/or fragmentation of a group of elastin sheets from the normal circumferential direction [(**Figure 1D-a’**, red arrows) and (**Figure 1D-b’** asterix), respectively]. A moth-eaten appearance was also observed in a group of elastin sheets (**Figure 1D-b’ and c’**). The lamellar unit in HCM was sparsely packed at regions with ECM components compared to controls, with unoccupied spaces and areas of randomly orientated collagen (**Figure 1D-b’-c’**).

### 2. Interlamellar space composition [SMCs and ECM]

#### 2.1. SMCs characteristics in HCM aorta

We next investigated the expression of markers pertaining to SMCs, the cell population occupying the MLU ^21^. Immunohistochemical analyses showed a significant reduction in the expression of alpha-smooth muscle actin (α-SMA), Smooth muscle myosin (SMM), calponin, smooth muscle 22 (SM22), and integrin beta-1 (ITGβ1), and a significant increase calponin expression in HCM-aortic walls compared to controls (**Figure 2A and supplementary Figure 3**).

**Figure 2:**
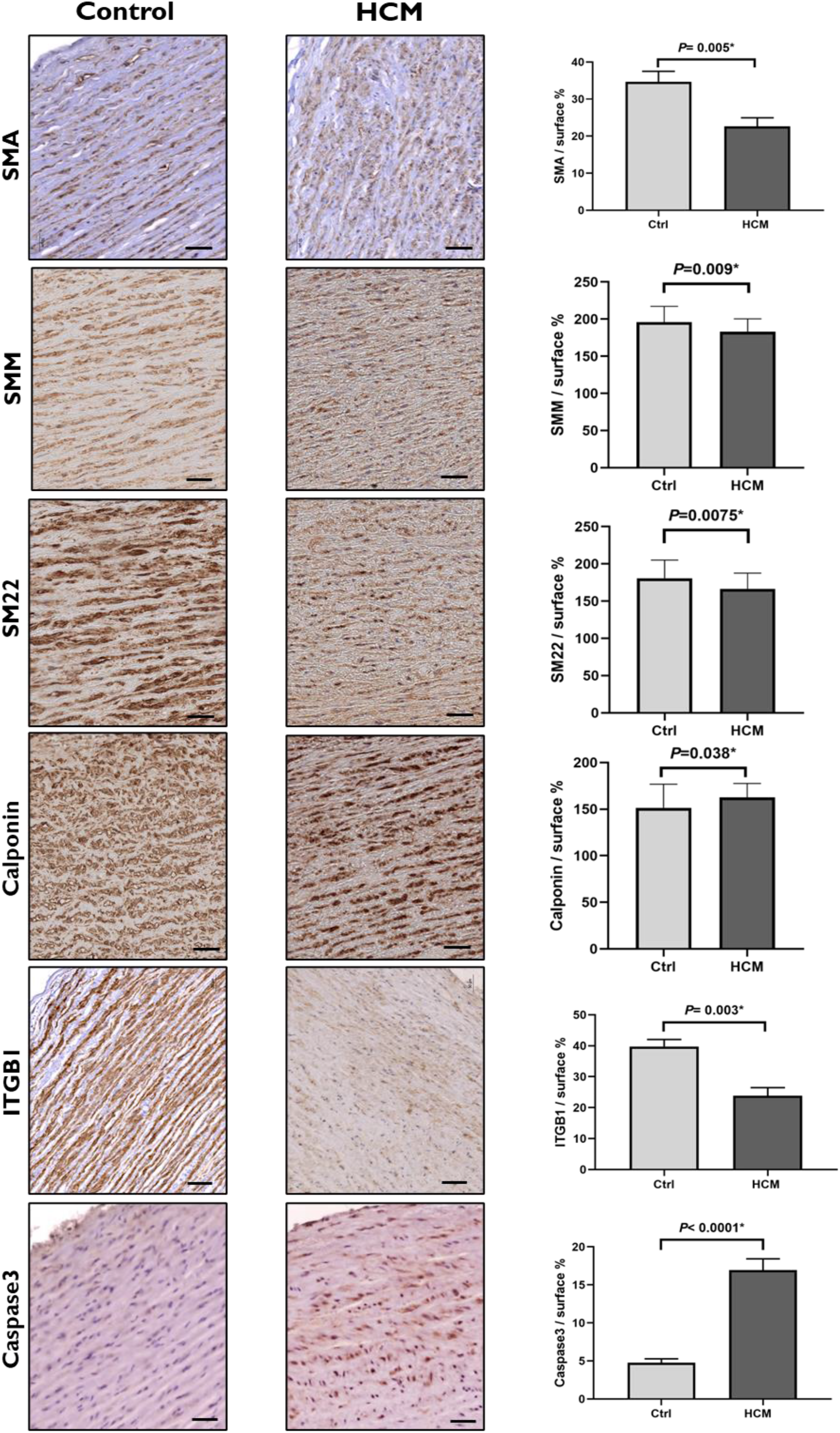
Expression of SMCs markers in HCM aortic wall. Immunohistochemical staining of α-SMA, SMM, SM22, Calponin, ITGβ1 and caspase3 in HCM-aortic biopsies (n=102) compared to controls (n=10). Scale bars are 200μm and 50μm. Bar graphs show a significant decrease of α-SMA, SMM, SM22 and ITGB1, and a significant increase of calponin and caspase 3, in HCM-aortic biopsies compared to controls.

To test whether the reduction in SMCs cells markers was associated with a depletion and/or apoptosis of SMCs, we assessed the expression of Caspase 3, as an apoptosis marker ^28^, which showed a significantly increased expression in HCM-aortic walls compared to controls (**Figure 2A**). Electron microscopy imaging showed that SMCs in HCM samples contained fat globules, vacuoles, condensed nuclei, and irregular cell outlines, which is indicative of necrotic changes compared to control SMCs. Nonetheless, nuclei of SMCs in HCM appeared to remain intact (**Figure 3**).

**Figure 3:**
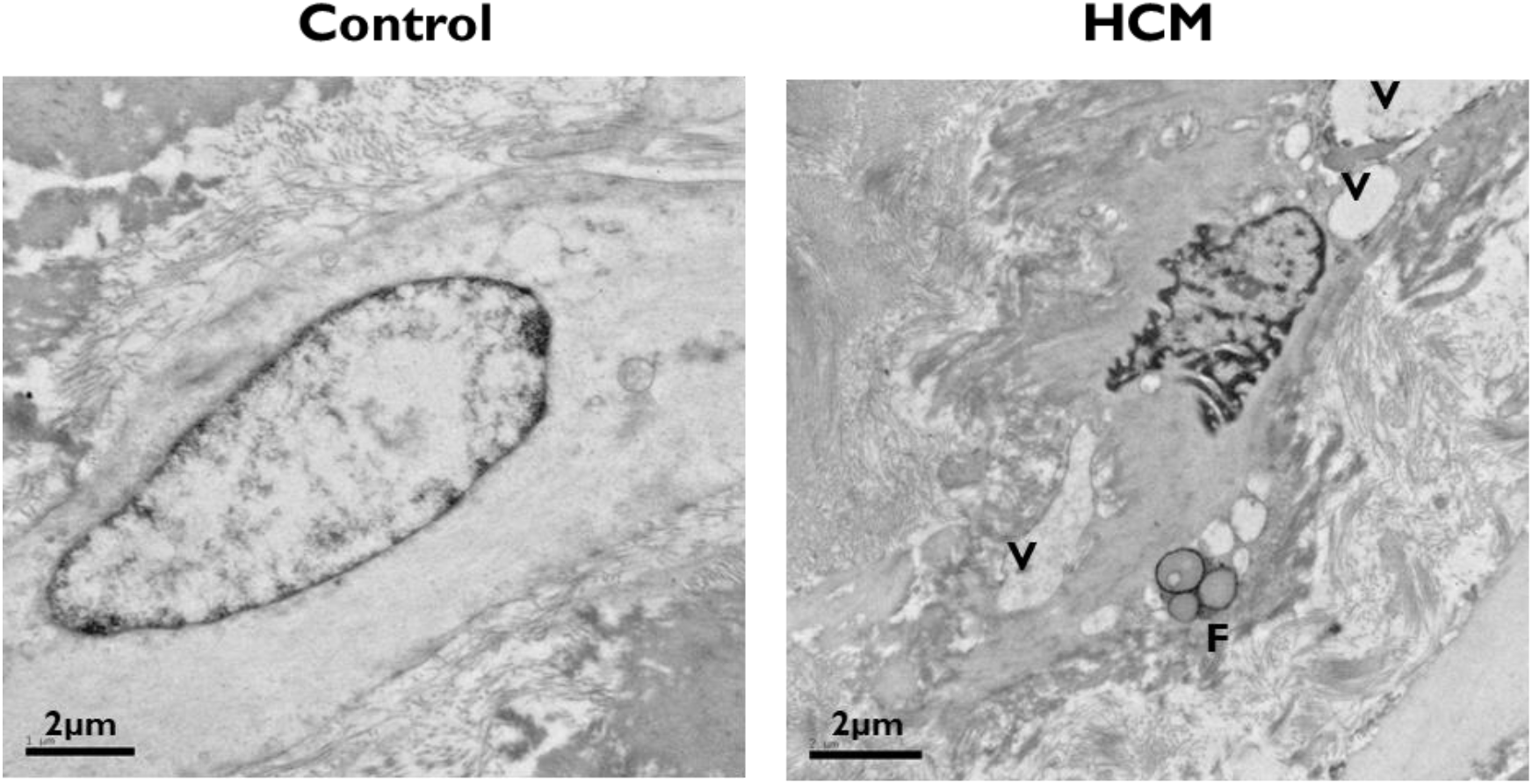
SMCs abnormalities in HCM aortic wall. Electron micrographs of control (n=3) and HCM (n=14) aortic walls, showing the altered structure in SMCs in HCM walls. F: fat globules and V: vacuoles

#### 2.2. ECM changes in HCM aorta – Collagen and Fibulins

Histopathological characterization of fibrous collagens in aortic wall tissues collected from HCM patients (n=102), showed a disorganization of collagen fibres compared to controls (n=10) (**Figure 4A**). Electron microscopy imaging of control collagen in transverse sections showed a minor variability in fibres diameter, however HCM samples showed a higher variability in collagen diameter and more loose fibres packing (**Figure 4B-a and b**). Similarly, longitudinal collagen fibres in control samples showed a minor variability in fibres thickness, whereas HCM samples had a group of thicker collagen strands with evident unravelling (**Figure 4B-c and d**, red arrow).

**Figure 4:**
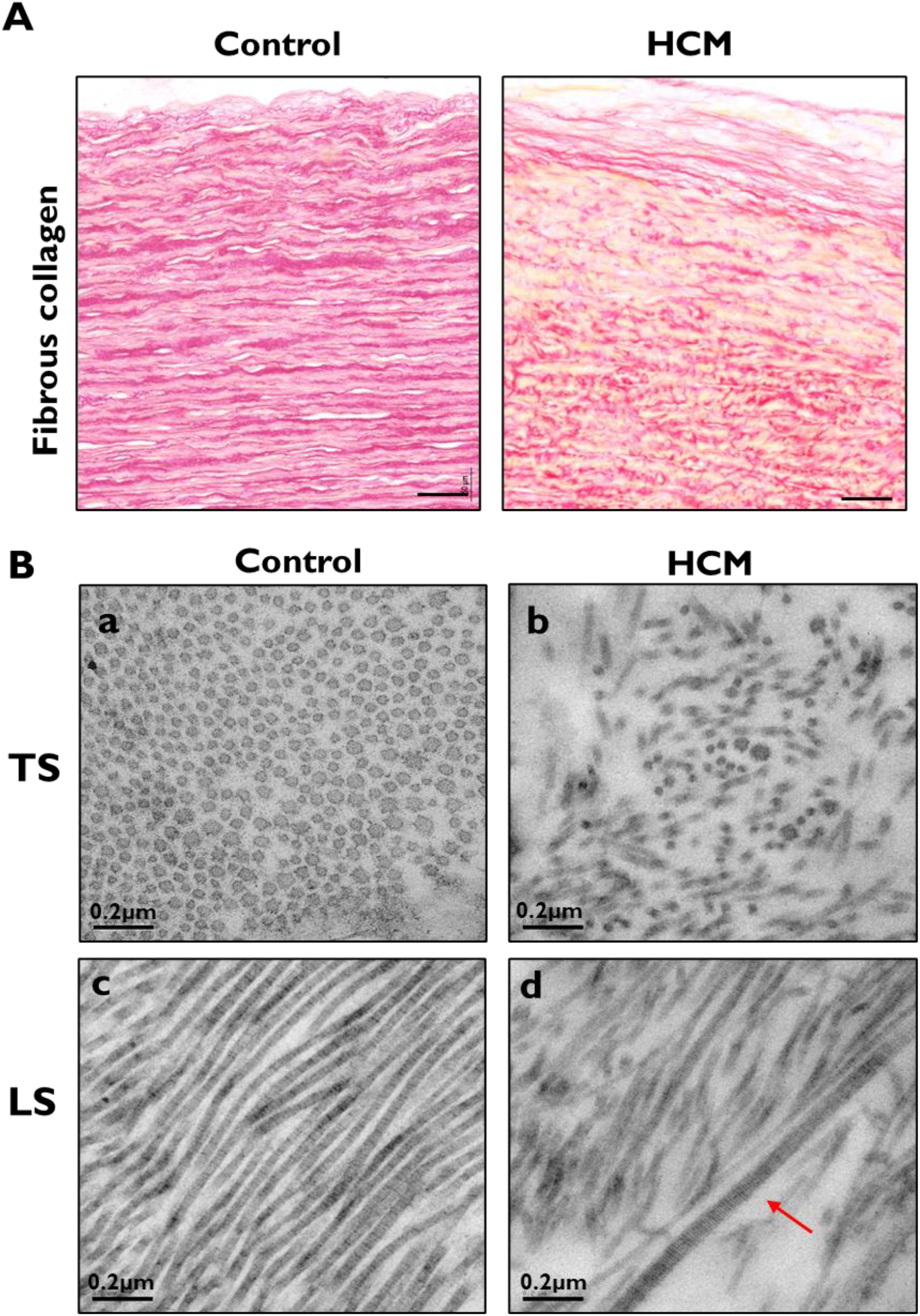
Collagen abnormalities in HCM aortic wall. **A:** Picrosirius red staining in HCM aortic biopsies (n=102) compared to controls (n=10). Scale bars are 50μm. **B:** Electron micrographs of control (a and b) (n=3) and HCM (c and d) (n=14) aortic walls. Scale bar are 0.2μm. LS: longitudinal section. TS: Transverse section.

In control aortic walls, ECM proteins, FBLN1, 2 and 5, were expressed in the elastin lamina in addition to the tunica media layer with a similar pattern to collagen and elastin fibres. In HCM-aortic walls, FBLN1 and FBLN2 expression was reduced, particularly in the tunica media, while FBLN5 was reduced across all wall layers, compared to controls **(Figure 5A)**. Immunoblotting analysis confirmed the reduced expression of the three FBLNs in HCM-aortic lysates (**Figure 5B and 5C**).

**Figure 5:**
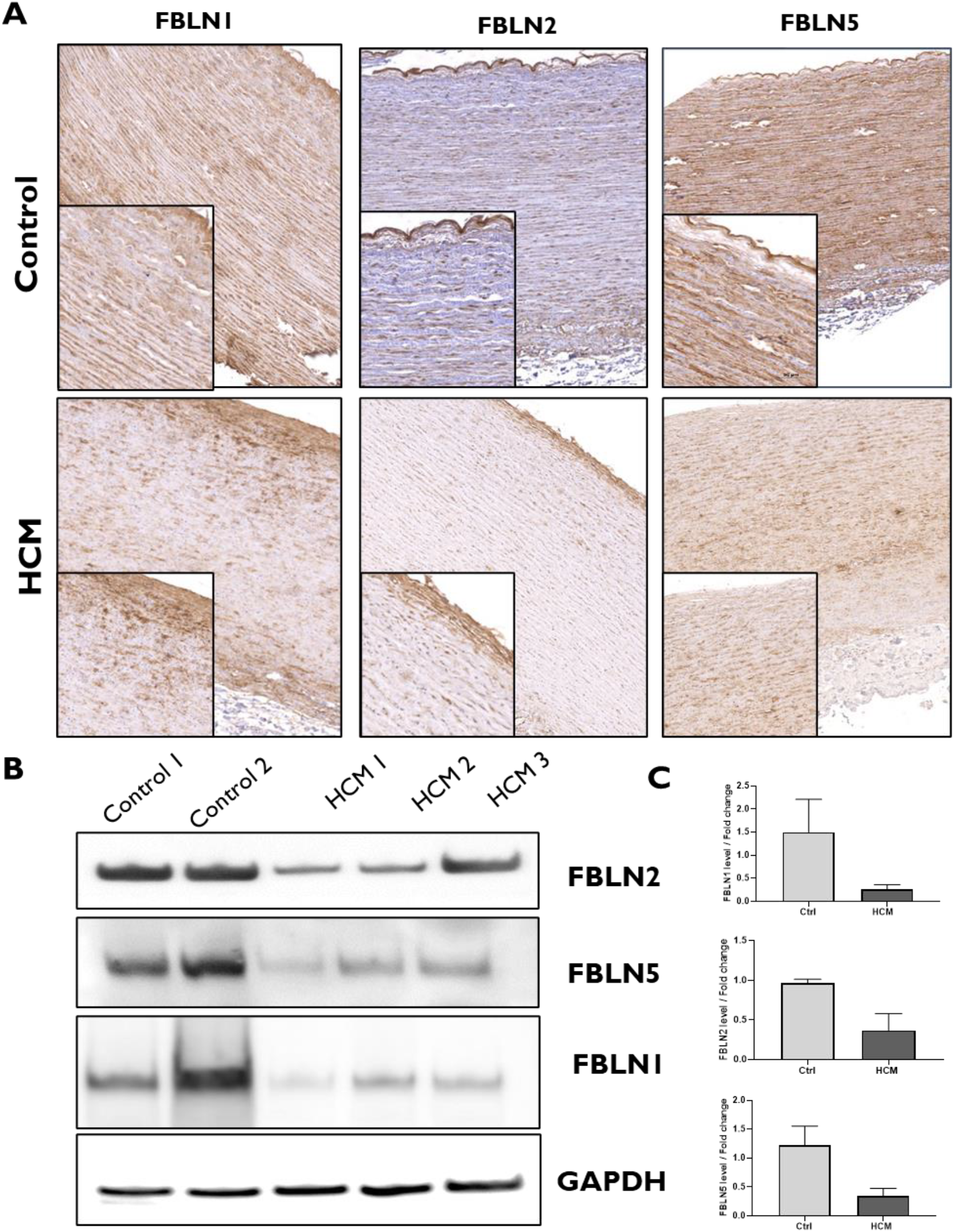
ECM abnormalities in HCM aortic wall. **A:** Immunohistochemical staining of FBLNs 1, 2 and 5 in HCM-aortic walls (n=102) compared to control walls (n=10). Scale bars are 200μm and 50μm. **B:** Immunoblot shows the reduction of total protein expression for FBLNs 1, 2 and 5 in HCM-aortic walls (n=6) compared to control walls (n=2). **C:** Quantification of FBLNs 1, 2 and 5 in HCM-aortic walls (n=6) compared to control walls (n=2).

## Discussion

This study shows significant changes in the medial lamellar unit (MLU) of the aortic wall in HOCM patients, associated with increased wall stiffness. Recent evidence has shown that HCM is not entirely due to abnormalities in the sarcomere ^2,3,7^. This notion has been supported by genome wide association studies (GWAS), which showed that HCM is linked to changes in genes involving many pathways in addition to the well-known variations in the sarcomeric genes ^7^.

One of the extracardiac manifestations of HCM is the altered stiffness of the aortic wall ^9,30–32^, and the systemic increase of pro-fibrotic markers ^33,34^. Aortic stiffness increases with age and is known to be an important prognostic indicator in patients with hypertension, atherosclerosis, and other diseases ^31,32,35^. In addition, aortic stiffness has recently been shown to correlate to exercise capacity in HCM patients ^11^. The current study confirms the increased stiffness of the aortic wall of HOCM patients, when compared to age-matched controls. To date, there have been no studies addressing the structural changes in the aortic wall of HOCM patients.

### Aortic Medial Lamellar Unit

The lamellar unit of the aorta represents the active core that plays a major part in determining the physical characteristics of the aortic wall ^15,18^. It was originally identified by Glagov and his colleagues, and has since been validated by many studies, that the number of the lamellae is related to the dimensions, tension, and pressure of the arteries ^15–17,27^. Recently, an advanced 3D modeling and imaging of the MLU showed two dense layers of elastin constituting the boundaries of the interlamellar space. The latter contains SMCs, oriented in a specific manner, as well as collagen fibres and other components of the ECM ^19^. Our study documents for the first time a decrease in the number of elastic lamellae and an increase in the interlamellar distance and composition. This could influence the elasticity of the arterial wall and increase its stiffness.

Electron microscopy and histological examination showed fragmentation in elastin sheets in HCM aortic walls. Elastin fibres confer compliance and recoil capacity to the aorta in response to mechanical stimuli ^13,36^. Elastin fragmentation is frequently observed in many aortopathies and has been shown to stimulate SMCs turnover ^23^, which further agrees with our analysis that identified a significant reduction in the surface and contractile markers of SMCs.

### The interlamellar space in HOCM

Apart from being wider in HOCM, the composition of the interlamellar space is altered. This is exemplified in the SMCs which showed changes in the orientation, shape, and size. These changes could possibly contribute to the increased stiffness by altering the vessel tone ^37,38^.

Our findings suggest that SMCs function could be further influenced by the reduction of ITGβ1 expression. ITGβ1 is a key signaling and mechano-transduction receptor of SMCs ^39^. We further show a significant reduction in the SMC contractile proteins which suggests the dedifferentiation of SMCs to a migratory and proliferative phenotype and/or activation of apoptosis ^40–42^. This agrees with the increased expression of caspase3 in HCM aortic walls, an observation that has been reported in many aortopathies, particularly aortic aneurysm ^43,44^. The significance and clinical relevance of these changes requires further investigation.

This study also identified changes in the interlamellar space, represented in collagen and ECM proteins (FBLNs). Physiologically, collagen fibres, particularly fibrous collagens I and III, provide tensile strength and overall rigidity to the aorta ^45,46^. Our data showed disorganized collagen fibres in HOCM aortas, which can further contribute to wall stiffness. FBLNs expression was reduced in HOCM aortic walls, particularly in the tunica intima and tunica media, suggesting their contribution to the lamellar unit’s conformation and function. This was associated with a reduction in the basement membrane protein, Collage IV (**Supplementary Figure 4**), which requires FBLN2 for proper assembly ^47^. FBLNs are a family of glycoproteins that have roles in ECM remodeling, tropoelastin maturation and collagen fibre assembly ^48–50^. A notion suggesting their relevance to the altered MLU.

Structural and functional data presented in this study highlight the importance of further unravelling the composition and role of the ECM and SMCs in the human aorta in both physiological and pathological conditions, as it relates to HOCM.

The limitation of this study is the fact that all studied patients had obstruction which could have influenced the results, however, the aortic biopsies were high up in the ascending aorta and it’s very unlikely to have been influenced by the disturbed pattern of blood flow in the aortic root.

In conclusion, these findings can be of special relevance to the management of HOCM by identifying high-risk patients. In addition, it could provide new therapeutic targets such as in the VANISH trial, which used Angiotensin-II receptor blocker-Valsartan that could be explained, at least in part, by the effect of the drug on the arterial wall ^51^.

## Data Availability

All underlying data are available in the article and in its online supplementary materials.

## Data availability statement

All underlying data are available in the article and in its online supplementary materials.

## Acknowledgement

The authors would like to thank Dr. Mona Allouba, Ms. Hadir Khedr, and Mr. Mohamed Elkhateb for their major contribution in acquiring the clinical data for control healthy cohort.

