## Supplementary materials for "Structural and Functional Characterization of the Aorta in Hypertrophic Obstructive Cardiomyopathy"

### Patients characteristics

HCM cohort included more males and were relatively older than the control group. 50% of HCM patients were incapacitated with dyspnea on less than the ordinary effort (NYHA class III). They had severe left ventricular hypertrophy, smaller LV cavity volumes throughout the cardiac cycle, high ejection fraction with severe LVOT obstruction and diastolic dysfunction, evidenced by cardiac imaging tools, Echocardiography, and cardiac MRI.

### Supplementary Figure 1

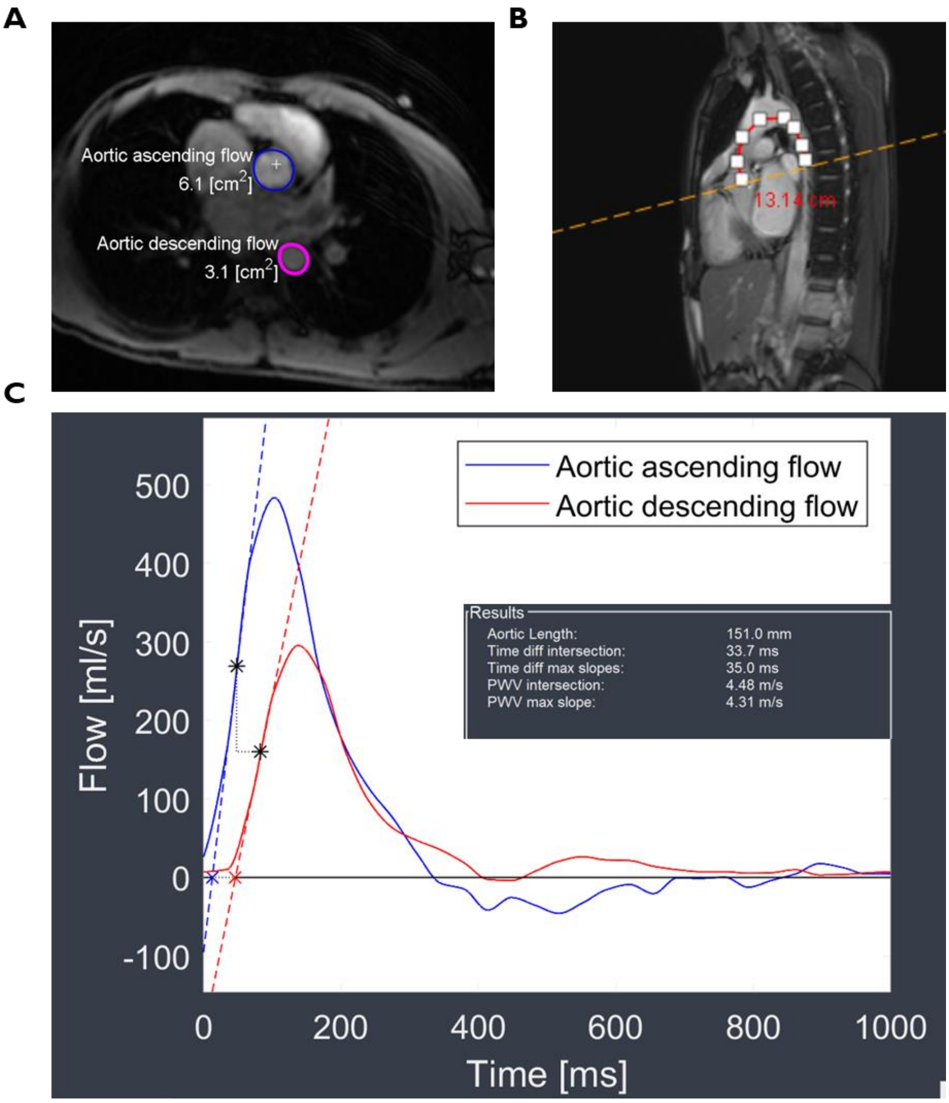


**Supplementary Figure 1: Measurement for Aortic pulse wave velocity (PWV)**. **A:** Phase-contrast cine transverse view. **B:** Aortic Sagittal view (localizer) to measurement of the transit distance in the aorta. **C:** Flow wave curves of ascending and descending aorta after peak flow normalization. Transit time is measured as an average time difference using the least squared estimates between all data points on the systolic upslope of the ascending and descending aortic flow curves.

### Supplementary Figure 2

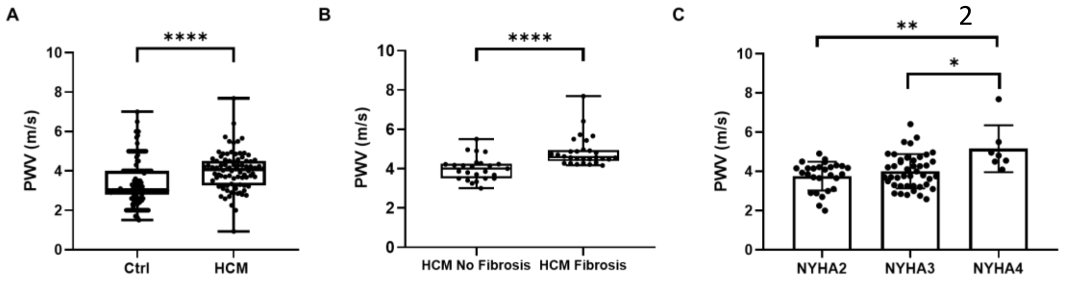


**Supplementary Figure 2: PWV in the aorta of HCM patients. A**: PWV was significantly higher in the aorta of HCM patients (n=86) (3.86 ±0.98 m/s) compared to healthy controls (n=166) (3.21 ±1.07 m/s) (P=0.001).**B:** Patients with reported myocardial fibrosis (assessed by CMR) (n=29) had a significantly higher PWV (4.85 ± 0.76 m/s) compared to those with no myocardial fibrosis (n=25) (4.01 ± 0.59 m/s) (p <0.0001). **C**: PWV levels in aortas of patients with NYHA class II (n = 37), class III (n = 44) and class IV (n = 7). The age range of NYHA IV patients [Mean 48.78 ± 8.09] was higher compared to NYHA II patients (Mean 40.13 ± 15.5) (p <0.01) and NYHA III patients (Mean 39.04 ± 12.9) (p = 0.0261).

### Supplementary Figure *3*

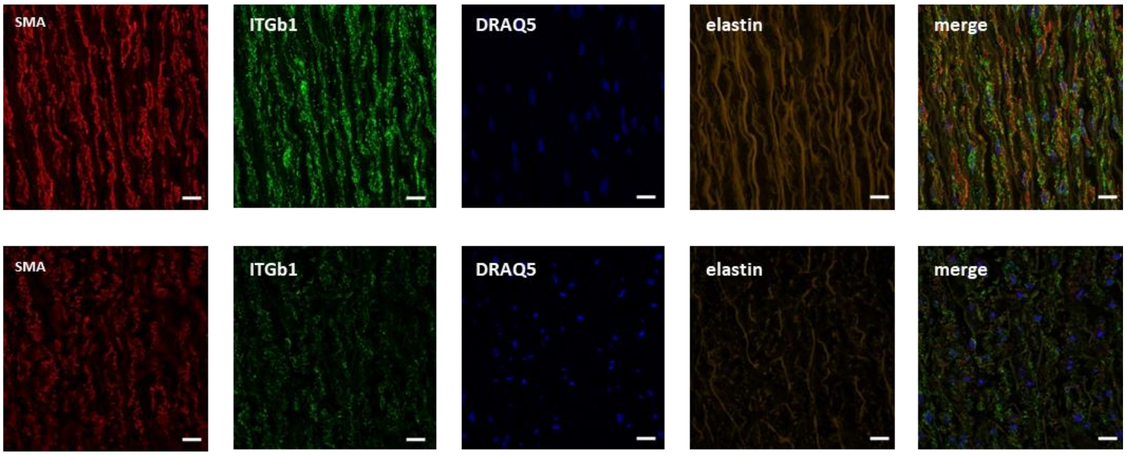


**Supplementary Figure *3*: Disruption in the organization of lamellae in HCM aortic walls.** Immunofluorescence-Confocal Microscopy of HCM aortic wall vs a control showing reduced expression of smooth muscle cells markers (SMA-red, ITGB1-green) in HCM aortic walls, associated with disorganization of the aortic lamellae. Scale bars are 50µm.

### Supplementary Figure 4

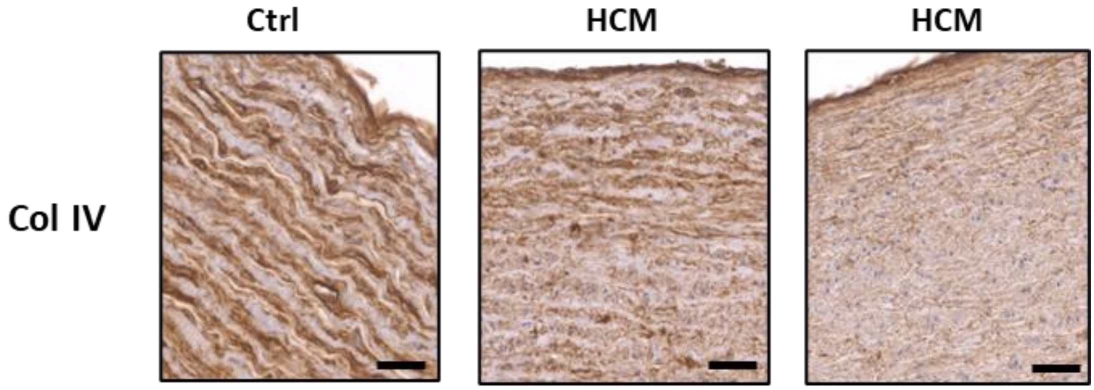


**Supplementary Figure 4: Expression of Collagen IV in HCM aortic walls.**

Immunohistochemical staining of Col IV in HCM aortic walls (n=12) compared to ctrl (n=3). Scale bars are 50µm.

### Supplementary Methods

#### Pulse Wave Velocity

The aortic path length was measured manually along the centre-line of the aorta in the oblique sagittal aorta images, between the acquired through flow planes in the aortic valve and the descending aorta. For quantitative flow curves, the flow curves from the aortic valve and the descending aorta at the level of the mid-thoracic were superimposed and intersecting tangents between upslope and baseline were determined. The time delay was calculated from the points at which these upslope tangents intersected the baseline tangents.
